# A Virtual Music Mindfulness Tool for Individuals of African descent during COVID-19

**DOI:** 10.1101/2024.12.23.24305623

**Authors:** Eghosa Igbinobaro, Amanda Watts, Oluwatofunmi Oshodi, Myrline Cleo Emile, Aldwin Soumare, Percy Takyi, Thema Haida, Hanifa Washington, AZA Stephen Allsop

## Abstract

Mental health disparities result from complex factors, including differential diagnoses, lack of access to standard mental health treatments, and inconsistent application of treatments when care is accessed. The COVID-19 pandemic exacerbated these disparities as marginalized groups had less access to testing and care while having higher infection rates. Community-based forms of care, such as music and mindfulness, are affordable and accessible options that can potentially address present mental health disparities. Specifically, music and mindfulness tools can be delivered virtually and are effective means for treating stress-related conditions. However, previous studies measuring the impact of music and mindfulness on stress-related conditions lack representation of people of African descent (PADs) and other marginalized communities. Among many reasons, bias and mistrust in recruitment strategies contribute to the lack of representation in these research studies. In this pilot study, we tested the feasibility of PAD recruitment in a virtual community-based music mindfulness research study and measured the impact of participation on perceived stress. Participants (14) with diagnosed anxiety were enrolled in an 8-week virtual music mindfulness program. We found a significant correlation between time engaged in our music mindfulness platform and lower perceived stress. Future work will evaluate the adoption and efficacy of these virtual strategies in communities when combined with and replacing standard interventions.

## Introduction

The healthcare system in the United States has been unable to meet the increasing demand of the mental health crisis, leading to an epidemic of undertreated mental health conditions, especially among groups vulnerable to stressors [1,2]. In this context, the COVID-19 pandemic created new mental health challenges, and based on data from prior epidemics, there will be a significant increase in the prevalence of anxiety, depression, substance use, and other mental health conditions after the initial stressors of the pandemic [3,4]. People of African descent (PADs) are particularly vulnerable and have been disproportionately affected by the compounding effects of COVID-19 due to higher uninsured rates and unequal access to testing [5].

PADs already experience higher rates of morbidity and mortality as a result of stress-related health conditions, including cardiovascular disease, adverse birth outcomes, and diabetes [6]. In addition, mental health vulnerability is increased in PADs in the United States due to the unequal effects of poverty, education, and criminal justice sentencing [7,8]. Race-related trauma and socioeconomic vulnerability further exacerbate these effects to worsen health in PADs in the United States. A lack of access to traditional clinical settings for mental health treatment combined with various forms of unethical experimentation and exploitation contribute to distrust in traditional clinical settings [9,10]. In contrast, community-based forms of care show potential in addressing health disparities for PADs. In order to implement new treatments, they need to be affordable and highly accessible. Two approaches that meet these criteria are music and mindfulness-based approaches.

Various meditation and mindfulness principles and practices have been adopted into psychiatry and show significant reductions in multiple negative dimensions of stress [11]. Mindfulness-Based Stress Reduction (MBSR) is one of the most popular and structured adaptations of mindfulness practices in psychiatry and yields significant positive effects on anxiety, depression, chronic pain, cancer, and other disorders [12–14]. More recently, short daily meditation practices using new meditators [15] have also shown improvement in mood and cognitive function. Similarly, new online mindfulness meditation interventions have recently demonstrated stress reduction and positive effects on mood and cognition [16,17]. However, these studies do not have an adequate representation of PADs or other marginalized groups and are not designed to be culturally relevant or community-based [18].

Previous studies that include diverse populations show that adapting MBSR protocols to be more culturally relevant yields greater program completion, positive effects on depression and rumination, and decreased post-traumatic symptom severity [19–21]. Nonetheless, recent studies that include short daily meditation and mindfulness studies exclude individuals with a psychiatric history [15], continue to underrepresent PADs and other vulnerable communities, and are not typically designed to be culturally relevant or community-based [6].

Music is ubiquitous and contains fundamental aspects of communication that are present across cultures and is thought to have evolved due to its social function [22,23]. Previous work has shown music to alleviate symptoms of anxiety, depression, and schizophrenia [24–28]. Music alone can relieve many symptoms of stress, but it is also a preferred and effective support for meditation and mindfulness [29,30]. Still, studies investigating therapeutic effects of practicing music-mindfulness for stress management in PADs and other Black, Indigenous, People of Color (BIPOC) communities are scarce [6,21,31,32]. Thus, we tested the feasibility of recruiting PADs to participate in a virtual community-based music mindfulness research study and measured the impact of a virtual music mindfulness tool on perceived stress. We hypothesized that music mindfulness would decrease perceived stress over 8 weeks. We focused on perceived stress, given its common relation to anxiety and depression, major contributors to mental health morbidity and mortality.

## Materials & Methods

### Study Strategy

This study was approved by the Yale University Institutional Review Board (HIC#2000028866). Possible study participants included men and women of African descent, aged 18 – 45, with symptoms of depression or anxiety, and had a therapist or outpatient provider. Participants were recruited through community email listservs and social media platforms like Instagram, Twitter, and Facebook. Recruitment materials were promoted on our lab Instagram page, our community partner One Village Healing’s page, and various public social media pages focused on mental health (@therapyforblackgirls, @blackgirlinom, @diveinwell). An informational video and poster were used as recruitment material, including a QR code, a number to text, and a link to the social media recruitment page (Supplemental Material). A Google form was linked to promotional material to screen and enroll participants. Screened participants were given oral and written consent information and provided oral consent for the study. Eligible participants were assigned an ID number to store and track outcomes confidentially without personal identifiers. During analysis, participant data were coded/de-identified. Eligible participants were given the following surveys: Perceived Stress Scale (PSS) [33], Beck’s Anxiety Index (BAI) [34], Experiences of Discrimination (EOD) [35], Social Connectedness Scale (SCS) [36] and Absorption in Music Scale (AIMS) [37]. Scores were entered in Qualtrics and submitted to study researchers on Day 1 of enrollment, 4 weeks, and 8 weeks after enrollment (Figure 1). The study was conducted from October 2020 – December 2021.

**Figure 1.**
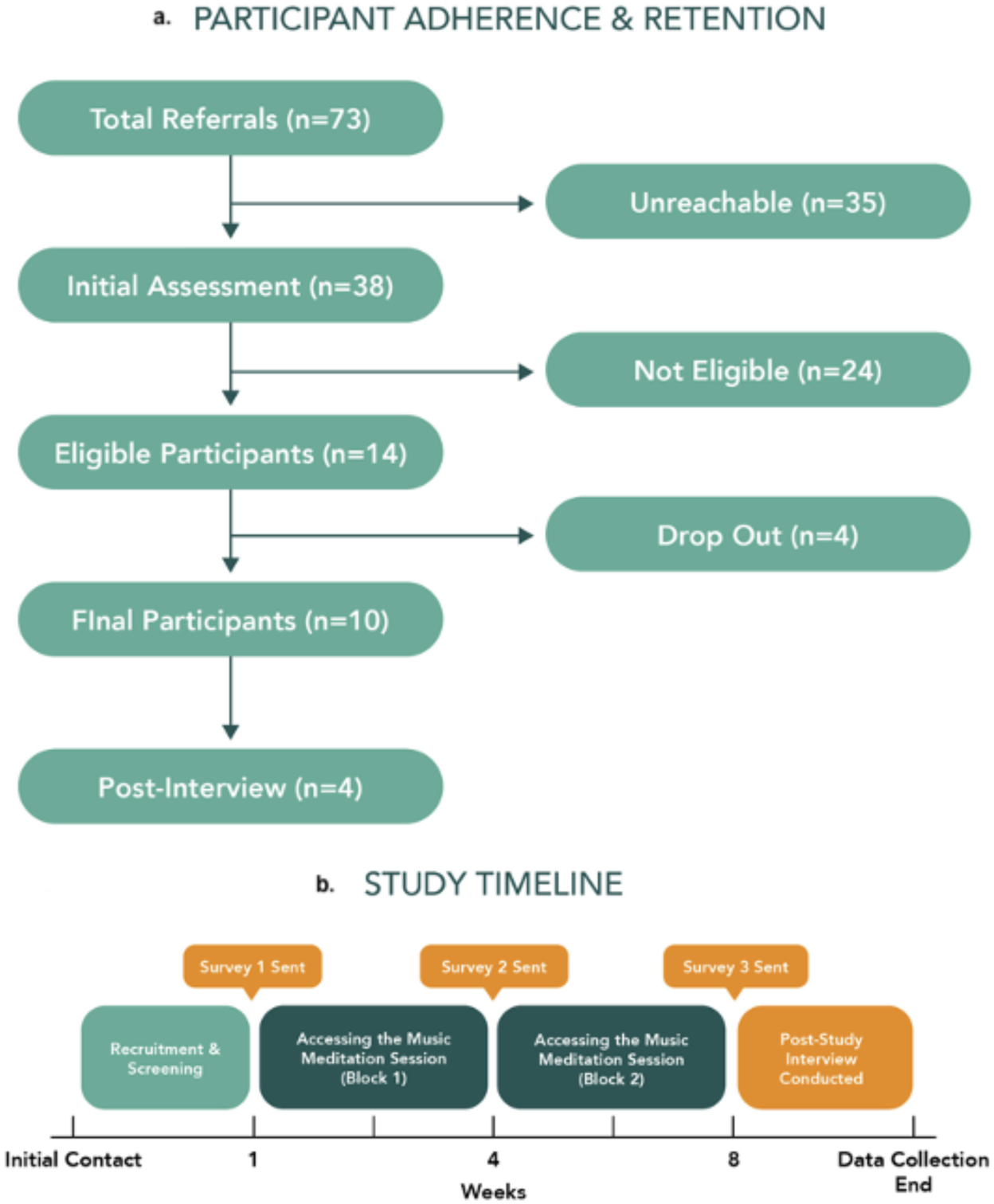
Study Recruitment and Timeline. **(a)** Participant adherence and retention, from referral to post-study interview. There were 10 final participants. **(b)** Study Timeline. Participants were given access to music mindfulness sessions for 8 weeks. They received surveys to collect data in Weeks 1, 4, and 8, and at a post-study interview.

### Music mindfulness sessions

Music for music mindfulness sessions was constructed based on principles of music preferred to support mindfulness [38]. A total of 7 instrumentals were created with tempos ranging from 70-100 beats per minute (**Figure 3**). Music was made in Logic Pro X with a Yamaha MOX6. The number of instruments for each session ranged from 5 - 8. Music was pre-selected by the principal investigator. Further details on the music used for each session can be found in Supplemental Materials. Mindfulness instructions were recorded over instrumental sessions and were aimed at fundamental meditation practices such as focus on breath, open awareness, and body scans (**Figure 3**). Vocals and instrumentals were mixed and mastered in Logic Pro X and uploaded to Wistia.com (Supplemental Material).

**Figure 2.**
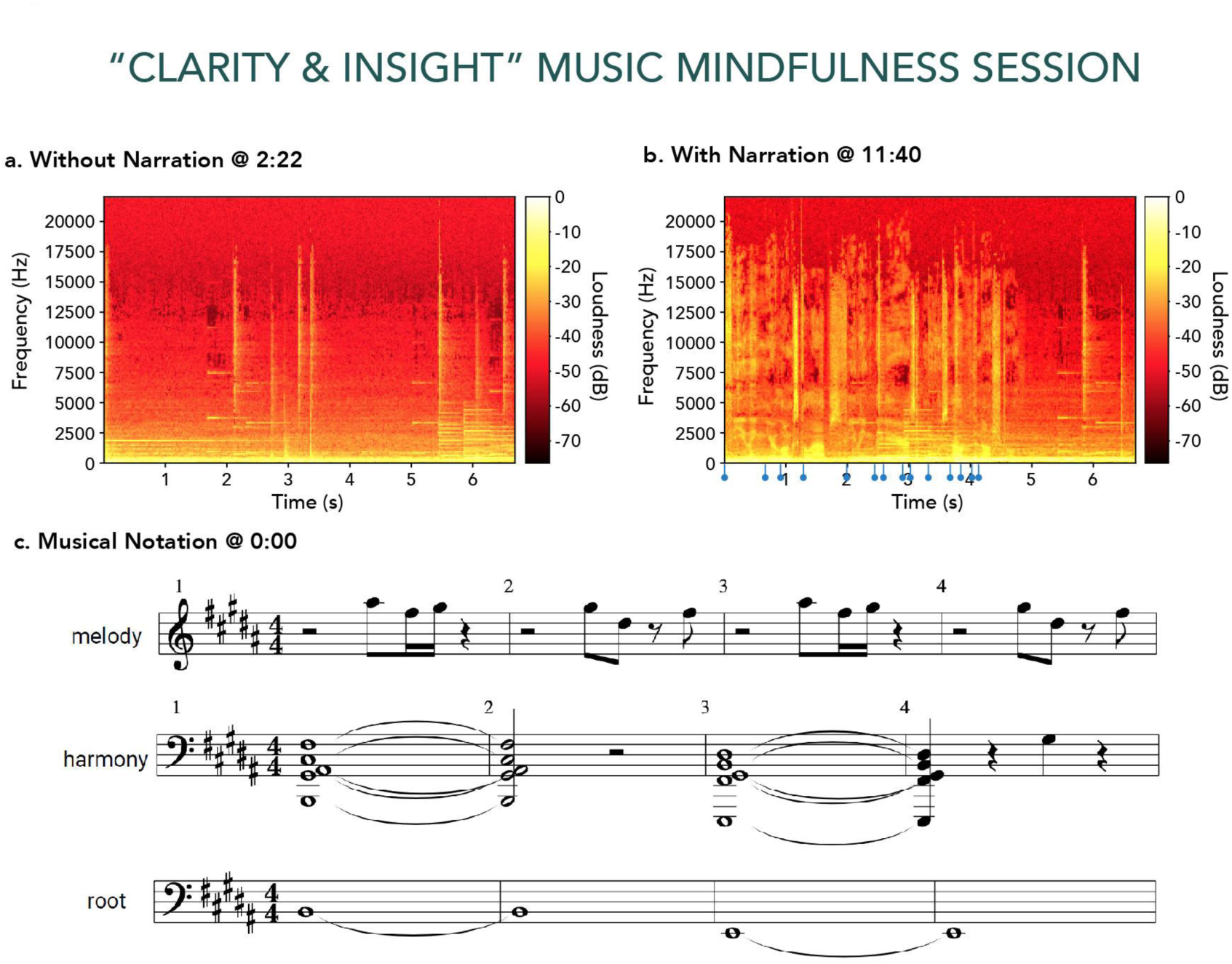
Physical and Musical Representations of Example Music Mindfulness Session. **(a)** spectrogram of sample without narration at 2:22 time point, **(b)** spectrogram of sample with narration at 11:40, and **(c)** musical notation for sample. Bright vertical bands (5000∼17500 Hz) are visible during seconds 2, 3, 5, and 6 of **(a)** and **(b),** corresponding respectively to notes in measures (bars) 1 and 2 of the melody in **(c)**. Bright horizontal bands ≤ 5000 Hz in **(a)** and **(b)**, respectively, denote the notes and corresponding harmonic overtones (resonant frequencies) present in the harmony and root notated in **(c)**. Horizontal bands representing resonant frequencies in the form of formants produced by the vocal tract of the narrator are also visible during the first 5 seconds of **(b)**. Blue tick markers on the time axis of **(b)** indicate the start of each word in the narrated phrase, “…feeling your lungs collapse, feeling the air as it moves out of the nostril.”

**Figure 3.**
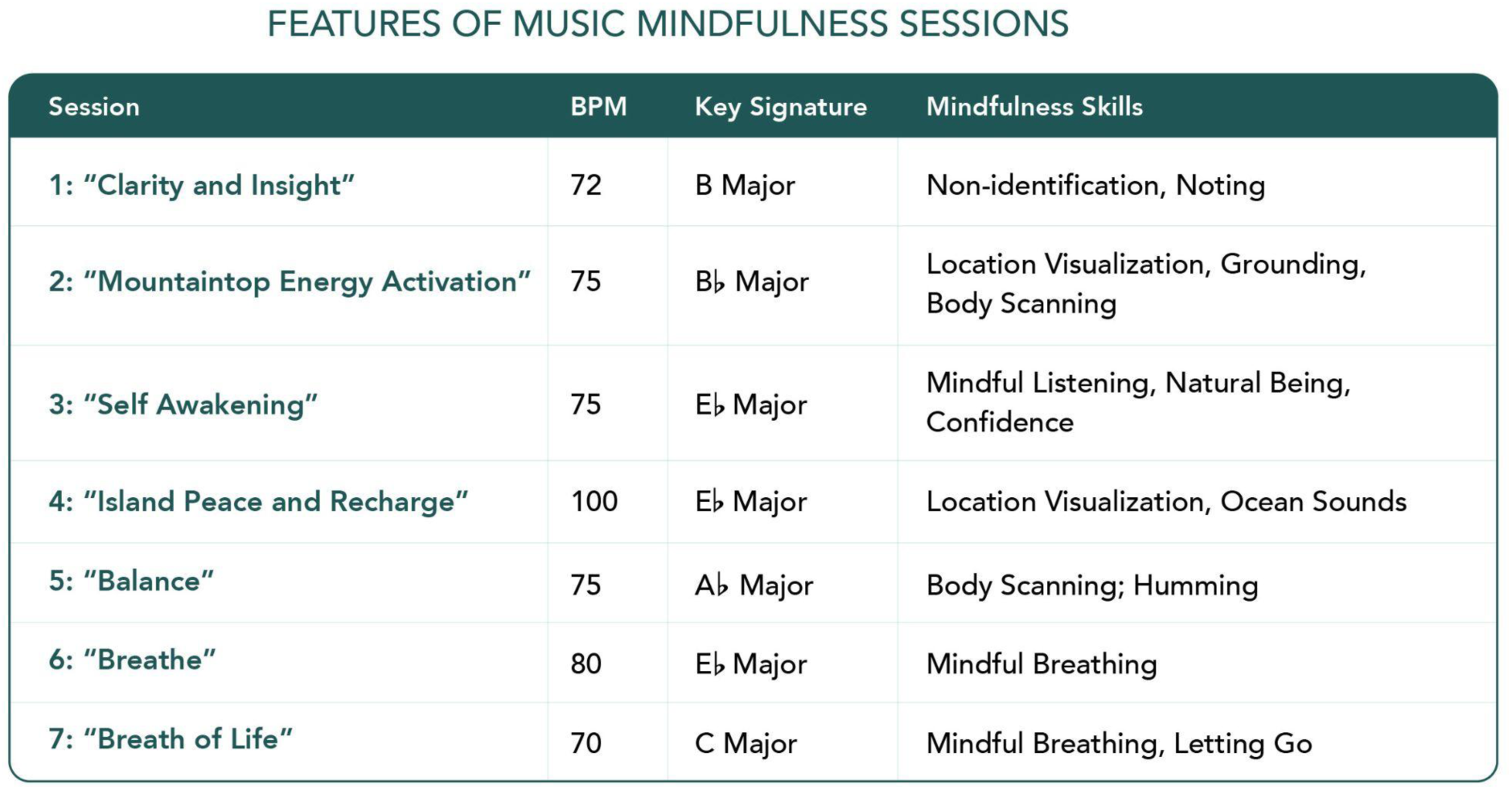
Features of Music Mindfulness Sessions. This qualitative table delineates seven distinct sessions, detailing their respective BPM (beats per minute), key signature, and associated mindfulness skill.

### Setting

On day 1, participants were given access to a virtual platform, Wistia.com, which hosted 15-minute pre-recorded, pre-selected music mindfulness sessions for daily at-home use via a computer or a mobile device. Participants also had access to weekly 1-hour virtual meditation classes through One Village Healing, a community organization located in New Haven, Connecticut.

### Participants

Participant eligibility was determined via phone screening, at which time consent to participate was also obtained (**Figure 1**). Participants were eligible based on the following criteria: a) African American/Person of African Descent with residency in the USA, b) age 18-65, c) moderate symptoms of anxiety or depression measured by the PHQ-9 and GAD-7, d) currently treated by a therapist or outpatient provider. Exclusion criteria included a) regular meditation practice of more than 20 minutes per week, b) has taken a meditation course or intervention in the past year, c) any hospitalizations in the past year, d) suicidal ideation, e) self-injurious behavior or homicidal ideation, f) audio-visual hallucinations or other signs of psychosis, g) diagnosis of a seizure disorder or other neurological disorder, h) history of thyroid or cardiovascular disease, i) active substance use disorder, j) current smoker, k) following medications: Benzodiazepines, Ambien, Intermezzo (zolpidem), Sonata, Rozerem (ramelteon), Notec, Precedex, Lunesta (eszopiclone), Trazodone, Seroquel, Remeron.

### Results

We implemented an inclusive recruitment strategy by creating culturally sensitive flyers and videos and sharing them via community-based listservs and social media platforms (Supplemental Material). Relatability, senses of empathy and collaboration with participants, and accessibility were conveyed through these materials by visually representing PADs in a variety of states. We also provided digestible statistics on mental health disparities and implemented other design and content strategies [39] (Supplemental Material).

A total of 73 participants were recruited for screening (**Figure 1**). Of those, we were able to contact 38 to complete the initial assessment. After screening for eligibility, 14 participants were enrolled. Ineligibility was mainly due to participants endorsing GAD7 and PHQ9 scores above the exclusionary threshold. Out of the 14 enrolled participants, 4 were lost to follow-up. A total of 10 (mean age = 37 ± 12.73 years; 26.3% of those who completed the initial assessment; Supplemental Material) were enrolled as participants, and 5 subjects completed all three surveys administered at Weeks 1, 4, and 8 (**Figure 1**). The average score reported via the AIMS did not significantly differ from normative data. The post-study interview was completed by 4 subjects (40% of participants).

Music mindfulness sessions were created using principles for music designed to support mindfulness. Evidence-based prompts were used to provide various mindfulness instructions [15,40–42]. For example, Session 1(“Clarity and Insight”) led listeners to employ mindfulness tools such as non-identification and noting [15] (Supplemental Material). This session was in the key of B Major at 72 beats per minute. Spectrograms and audio forms of sections display the difference in frequency and loudness during the non-narrated (**Figure 2A**) and narrated portion of (**Figure 2B**) sessions. The corresponding musical score provides the written notation for the spectrograms (**Figure 2C**).

Participants listened to an average of 41.95 minutes (SD = 39.01 min) of all seven music mindfulness sessions (**Figure 4**). Reported perceived stress (via PSS; mean baseline score = 16.5 ± 5.6) and anxiety (via BAI) were compared to listening during study Block 1 (Weeks 1 through 4) and 2 (Weeks 4 through 8). An inversely correlated trend was observed between reported perceived stress and listening time during Blocks 1 and 2 (Block 1: Pearson’s r = -0.7680, p < 0.05 (=0.04373); Block 2: Pearson’s r = -0.7042, p > 0.05 (=0.1183)) (**Figure 4 and 5**). An inversely correlated trend was also observed between reported anxiety and listening time during Blocks 1 and 2 (Block 1: Pearson’s r = -0.3748, p > 0.05 (=0.4075); Block 2: Pearson’s r = - 0.5854, p > 0.05 (=0.2222)) (**Figure 5**).

**Figure 4.**
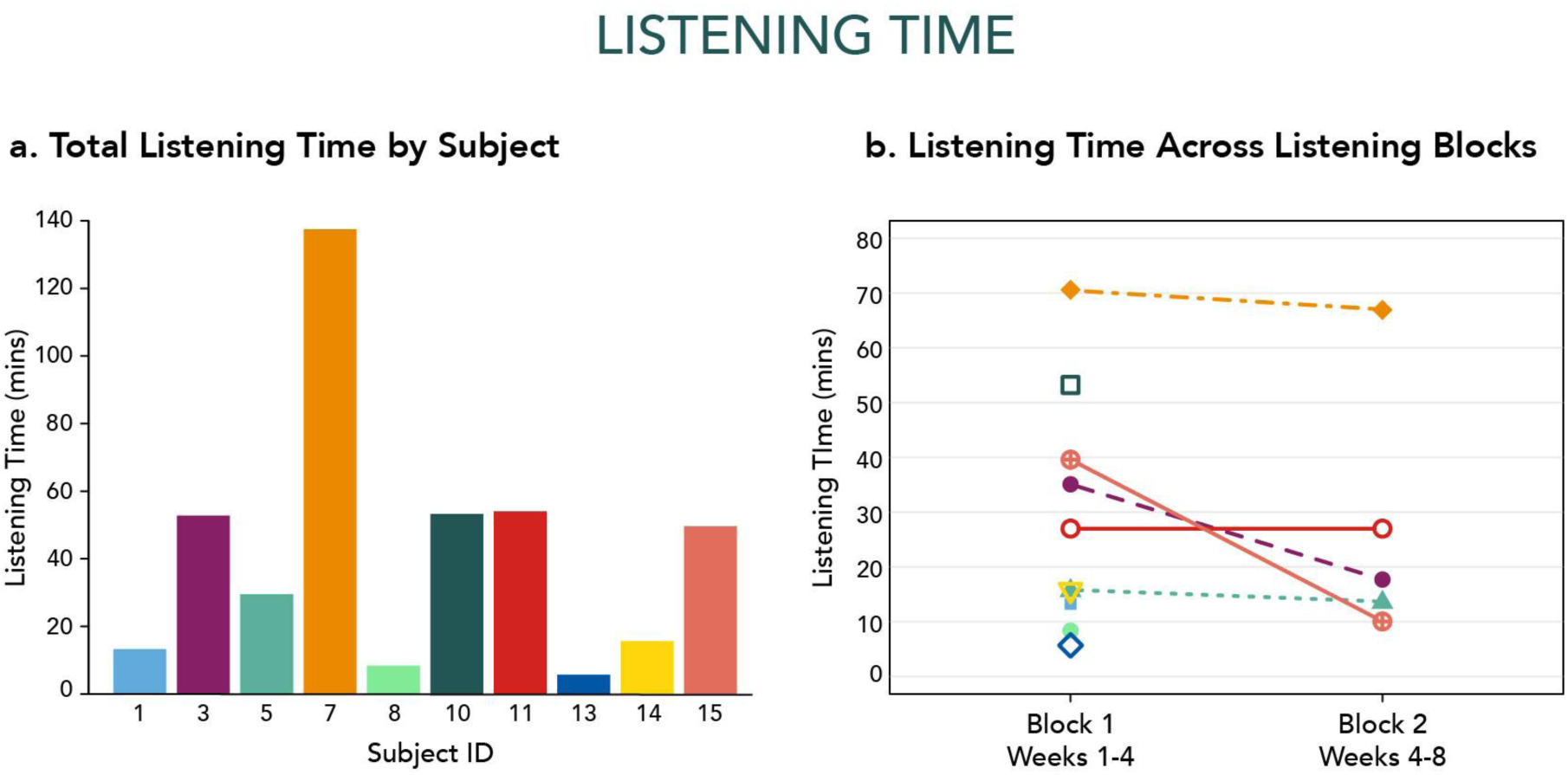
Music Meditation Video Listening Time. **(a)** Total listening time by subject (mean listening time: 41.95 ± 39.01 min). **(b)** Listening time by subject across listening blocks. Listening Block 1 corresponds to Week 1 to Week 4. Listening Block 2 corresponds to Week 4 to Week 8. Each color represents one participant across graphs. Closed-shaped points indicate participants who completed all three surveys (administered during Weeks 1, 4, and 8) while open-shaped points denote subjects who completed less than three surveys.

**Figure 5.**
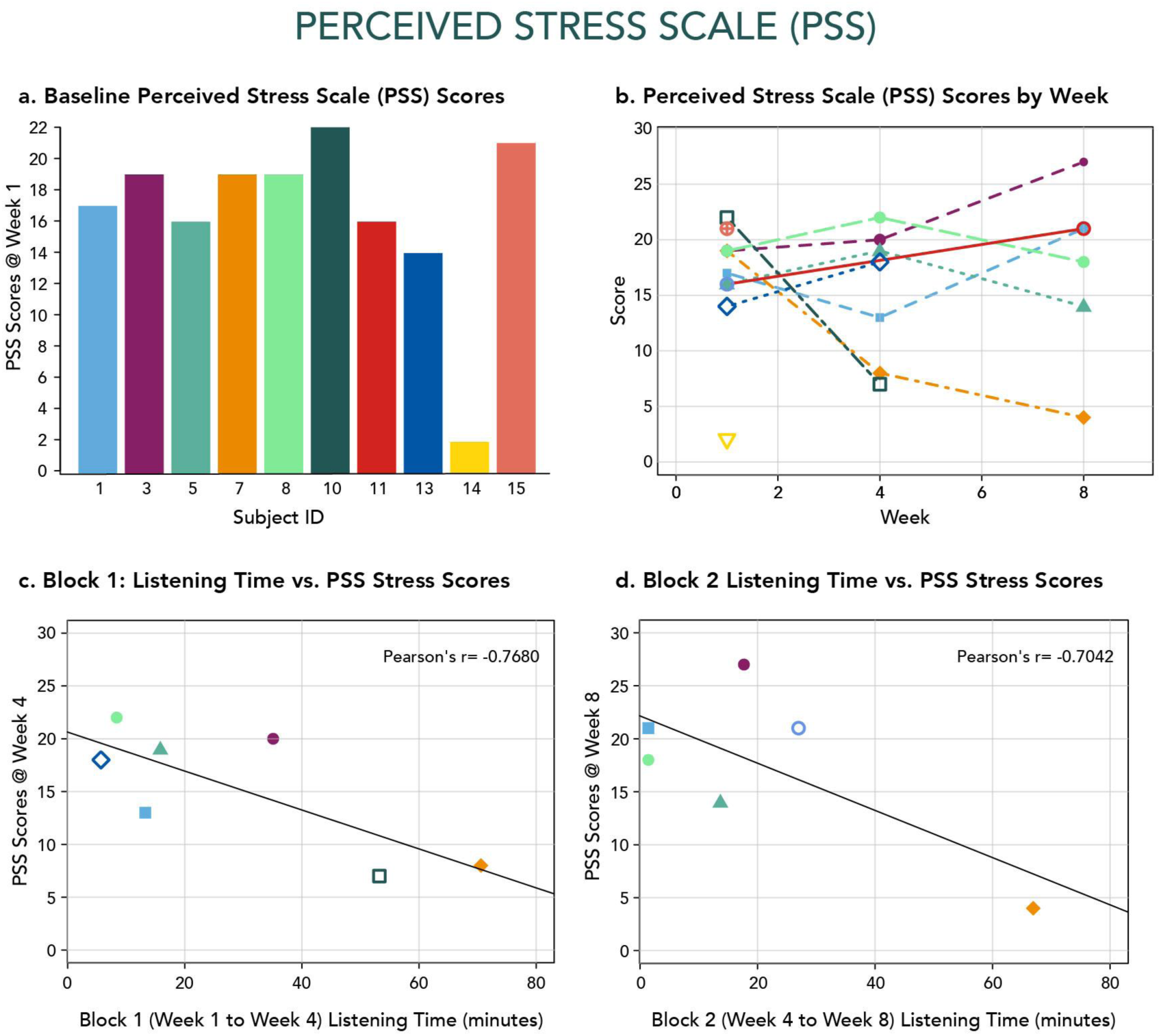
Perceived Stress Scale (PSS) scores by subject. Inversely-correlated trend observed between reported perceived stress and listening time. **(a)** PSS scores collected at Week 1 (mean PSS score: 16.5 ± 5.6). **(b)** PSS scores collected at Weeks 1, 4, and 8. **(c)** PSS scores collected at Week 4 versus listening time during Block 1 (Weeks 1 through 4); Pearson’s *r* = -0.7680, *p* < 0.05 (=0.04373). **(d)** PSS scores collected at Week 8 versus listening time during Block 2 (Weeks 4 through 8); Pearson’s *r* = -0.7042, *p* > 0.05 (=0.1183). Each color represents one participant across all graphs. Filled points indicate participants who completed all 3 surveys (administered during Weeks 1, 4, and 8) while open points denote subjects who completed less than 3 surveys.

Post-study interviews revealed that participants continued to use the techniques they learned in the study in their everyday lives, citing meditation, breathing, and humming exercises as some tools used. We also asked post-interview respondents to reflect on how their understanding of mindfulness changed as a result of participating in the study. Subject answers included recognizing the “powerful impact” of mindfulness and “the importance of ritual/routine for wellness”. Subjects also endorsed that mindfulness practiced during the study “formed the basis of [their] new healthy habit” and led them to be “more open” to the practice. An acknowledgment of not continuing to practice daily and a need “to become more disciplined in order to get the full benefits of the practice” was also stated.

## Discussion

As the mental health crisis worsens, communities of color will continue to suffer from mental health disparities. While there are therapies available to help with mental health, they need to be more affordable and accessible. One solution is to use music and mindfulness techniques, which can rely on virtual platforms, to deliver evidence-based tools that are affordable and accessible. Here, we designed and delivered a virtual music mindfulness therapy to address mental health symptoms. To test its feasibility in a marginalized population, we recruited people of African descent to participate in a community-based music mindfulness research study and measured the impact of this tool on perceived stress. While our feasibility data are preliminary, we were able to successfully demonstrate that PADs will participate in music mindfulness research through a community-based, social media approach. Interestingly, all of our enrolled participants were self-identified women; thus, research studies that include music and mindfulness may be particularly useful for addressing disparities in women’s health. Future work will be aimed at targeting male participants through community organizations that focus on men’s health. Of note, some potential participants, including men, were not eligible after screening for various reasons, including age and low or very high GAD7 or PHQ9 scores.

Post-study interviews with participants suggest the value of music as a means of attracting community members to participate in potential research or treatment. Participants subjectively reported music as an important reason for interest in the study and found music mindfulness sessions helpful for symptoms of stress.

Despite our ability to engage community members initially, we did note a loss of contact with some after screening. We also observed that participants decreased engagement with our platform over the 8-week enrollment period. One strategy to address this in future studies is to continue outreach once participants are enrolled. Study team engagement with participants once enrolled could improve the utilization of music mindfulness sessions.

Our pilot data suggests a correlation between increased time engaged in listening to music mindfulness platforms and decreased scores on the perceived stress scale. This supports our hypothesis that music mindfulness engagement over 4-8 weeks will decrease perceived stress. Future studies will be powered to determine the efficacy of these music mindfulness sessions on perceived stress. In our study, we also found that simply engaging in listening to the sessions did not have an impact on social connectedness. Listening in group sessions could help to increase social connectedness as music has been implicated in driving social connection and pro-social behavior.

Lastly, music and mindfulness are core components of psychedelic therapy treatment [43–45]. Music has been a fundamental aspect of both set and setting for psychedelic studies for MDMA in PTSD and psilocybin in depression. As psychedelic therapy becomes more inclusive and accessible, new and diverse ways of incorporating music into psychedelic therapies will be needed [46]. Our pilot data suggests that culturally relevant music mindfulness tools may be potential avenues for expanding therapeutic accessibility and inclusivity. In addition, our work in the community suggests that using music-based protocols could help enhance inclusive recruitment strategies for psychedelic research studies and treatment [47].

Together, virtual tools that allow for easy mobile access to music mindfulness sessions may improve perceived stress which has important implications for anxiety, depression, and PTSD. Delivery of technology-driven, evidence-based treatments has already been successfully implemented in the Black church [48]. This may prove a promising, easily discernible strategy to provide evidence-based interventions to an underserved and undertreated population. Continuous collaboration with community partners in larger, controlled studies will evaluate the effectiveness of this music mindfulness tool in diverse community settings.

## Data Availability

All data produced in the present study are available upon request to the authors

## Acknowledgments

We want to thank Dr. Ayana Jordan for direction and feedback on community-based participatory approaches for recruitment and enrollment of minoritized populations. Thank you to Stellate Communications for further edits and manuscript preparation.

## Author Contributions

ASA was responsible for conceptualization and supervision. ASA, EI, and OO designed the study methods and protocols. EI, OO, MCE, AS, PT, AW, TH, and HW performed participant screening, consent, enrollment, and data acquisition. AW and ASA performed data analysis and interpretation. AS, AW, EI performed software setup and maintenance. ASA, EI, OO, and AW wrote the initial draft of the manuscript. ASA, EI, OO created drafts, critically reviewed and edited drafts, and provided final approval of the manuscript. The authors agree on the accountability for all aspects of the work and the accuracy of the manuscript is met.

## Supplemental Materials

**Supplemental Figure 1.**
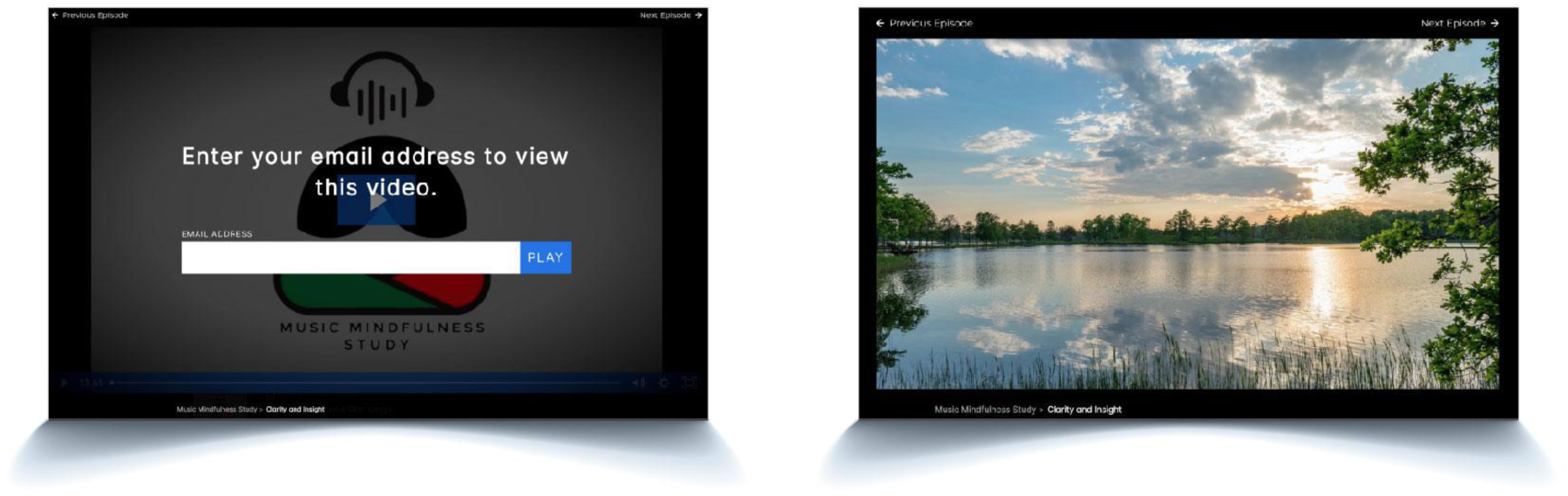
Virtual platform for music mindfulness sessions. Participants could access virtual sessions through wistia.com platform on a desktop, laptop, or mobile device. To listen to a session, participants entered their email addresses, allowing access for us to track viewing occurrences and time over the course of the study.

**Supplemental Figure 2.**
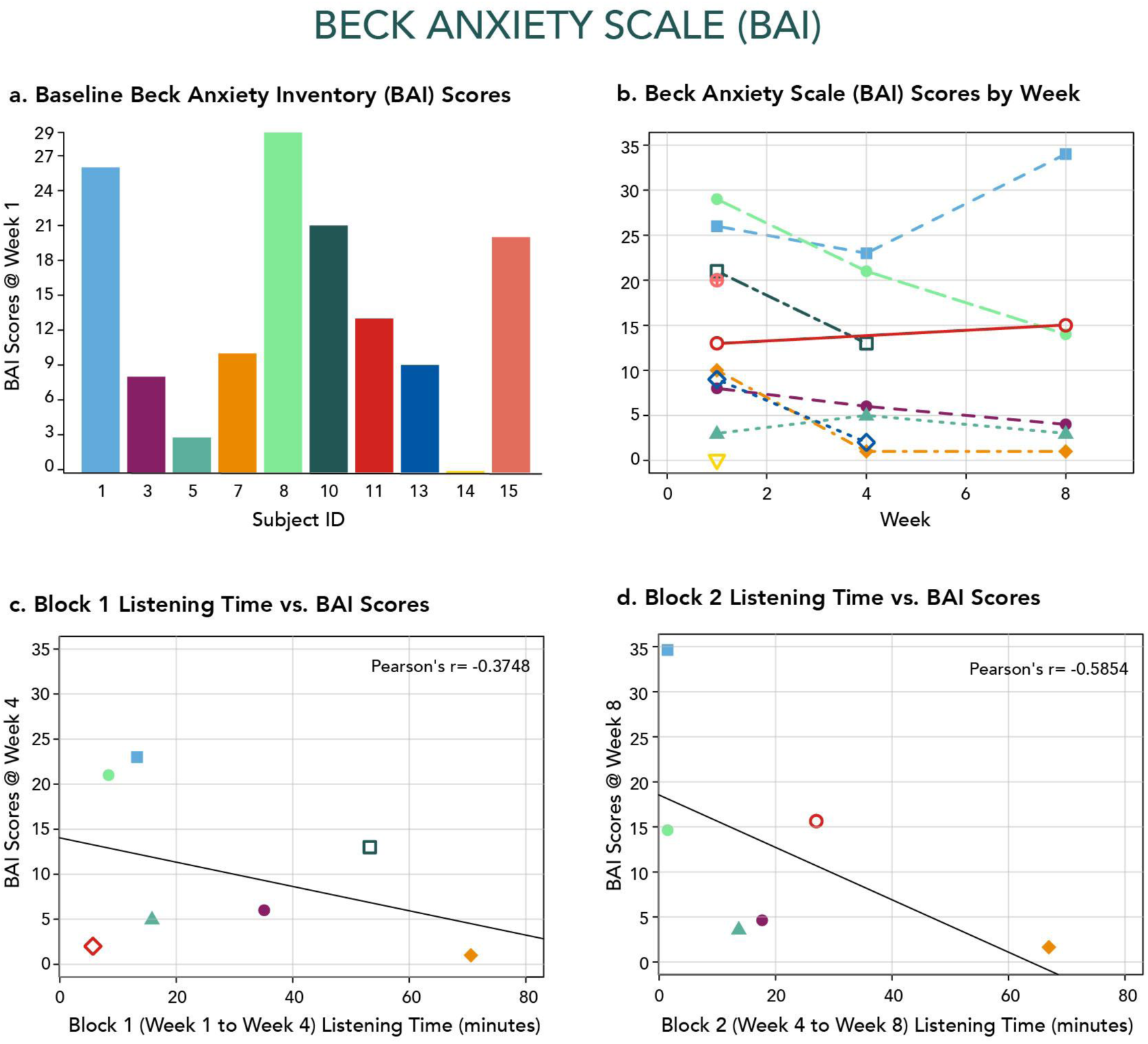
Beck Anxiety Inventory (BAI) scores by subject. Inversely - correlated trend was observed between reported anxiety and listening time. **(a)** BAI scores collected at Week 1. **(b)** BAI scores collected at Weeks 1, 4, and 8. **(c)** BAI scores collected at Week 4 versus listening time during block 1 (Weeks 1 through 4); Pearson’s *r* = -0.3748, *p* > 0.05 (=0.4075). **(d)** BAI scores collected at Week 8 versus listening time during block 2 (Weeks 4 through 8); Pearson’s *r* = -0.5854, *p* > 0.05 (=0.2222). Each color represents one participant across all graphs. Closed-shaped points indicate participants who completed all three surveys (administered during Weeks 1, 4, and 8), while open-shaped points denote subjects who completed less than three surveys.

**Supplemental Table 1.** Participant Characteristics.

| Participants | Number (%) |
| --- | --- |
| **Age**<br>M (SD)<br>18-28<br>29-38<br>39-48 | 37 (12.73)<br>4 (40%)<br>4 (40%)<br>2 (20%) |
| **Gender**<br>Female | 10 (100%) |
| **Race**<br>Black | 10 (100%) |
| **Ethnicity**<br>Latina | 2 (20%) |
| **Education (completed)**<br>Bachelor's<br>Earned at HBCU<br>Master's<br>Earned at HBCU<br>Professional | 4 (40%)<br>1 (10%)<br>3 (30%)<br>1 (10%)<br>3 (30%) |
| **Absorption in Music**<br>Average Baseline<br>(Week 1) Score | **M (SD)**<br>116 (33.5); norms:<br>113.5 (23.8) |

**Supplemental Table 2.** An Inclusive Media Strategy via Social Media for Participant Recruitment.

| Purpose/Aim | Recruitment media aspect |
| --- | --- |
| **Relatability** | **Video**<br>• Black person's hand drawing cartoons of black folk<br>• Black feminine-presenting voice narrating<br>**Video/Flyer**<br>• Showing PADs not only in distress, but also at peace and joyful |
| **Approachability** | **Video**<br>• Energetic Hip-Hop/R&B music in background<br>• Drawing of AZA, Principal Investigator, and T.wav\$, NtheLab Producer |
| **Credibility** | **Video/Flyer**<br>• Mention of affiliated institution and funding source |
| **Sensitivity** | **Not addressed** |
| **Accessibility** | **Video/Flyer**<br>• Multiple sign-up methods including QR code scanning, text messaging, and reaching out via Instagram, Twitter, and email |
| **Empathy/Common concern** | **Video/Instagram Posts**<br>• Inquiry of experience of mental health issues and assurance that viewer "is not alone" in this experience<br>• Acknowledgement of barriers to receiving treatment and inclusion in effective medical research<br>• Acknowledgement and citing of statistics on mental health disparities linked to race-related trauma and socioeconomic vulnerability experienced by PADs |
| **Collaboration/co-construction with participants** | **Video**<br>• Reference to community of PADs as "our community"<br>**Video/Flyer**<br>• Acknowledgement of participants' contribution to black mental health research through participation |
| **Transparency** | **Instagram Posts**<br>• "Meet the Lab" posts introducing public to lab members"<br>• "Music Mindfulness Minute": 1-minute reels with embedded audio samples of music mindfulness sessions |
| **Possible direct benefits** | **Video**<br>• Mention of benefits of engaging in music and mindfulness programs<br>• Mention of access of free mindfulness classes and music guided meditations to help manage stress |

**Supplemental Table 3.**
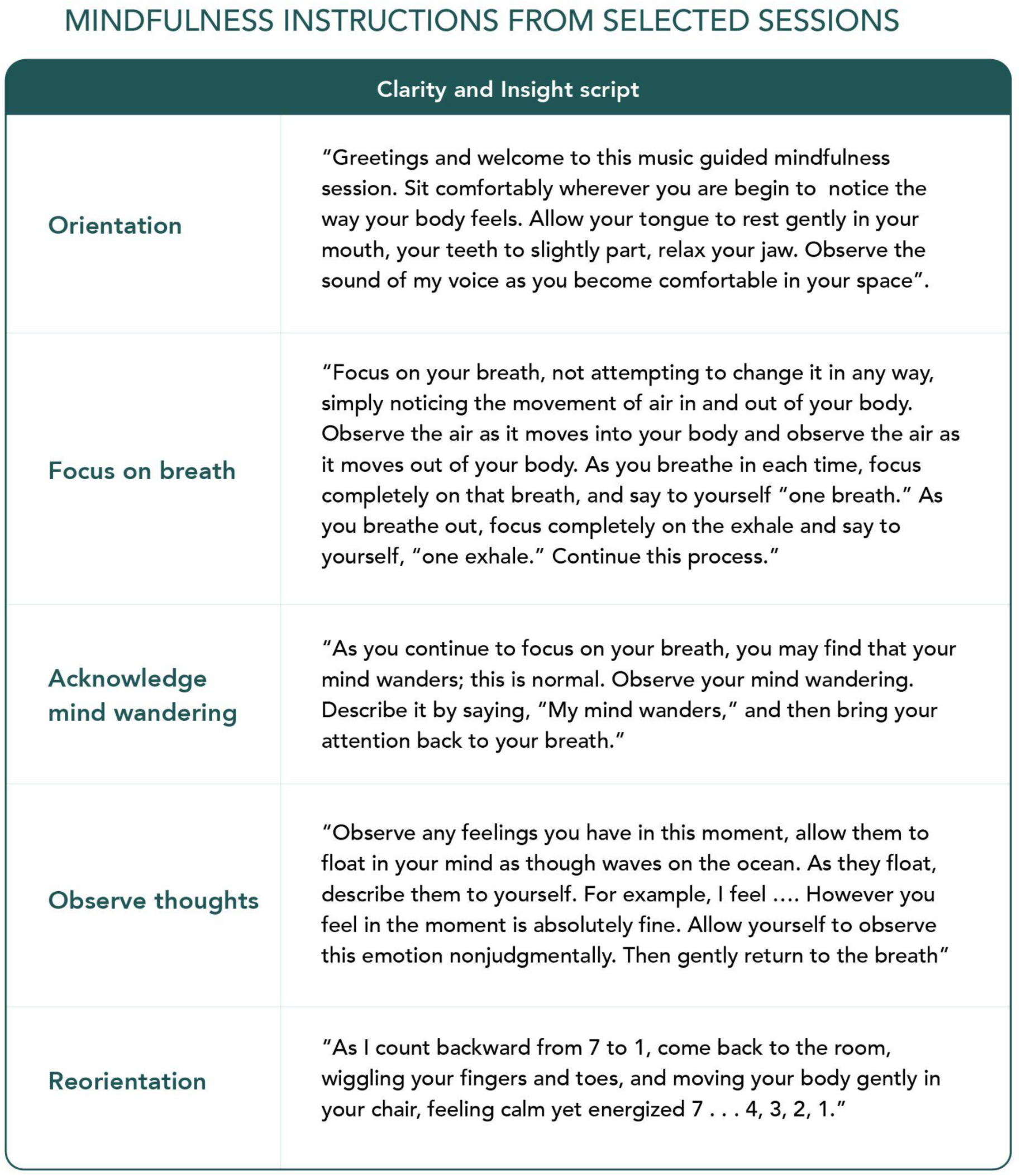
Mindfulness instructions from selected session.

| Clarity and Insight script |  |
| --- | --- |
| **Orientation** | "Greetings and welcome to this music guided mindfulness session. Sit comfortably wherever you are begin to notice the way your body feels. Allow your tongue to rest gently in your mouth, your teeth to slightly part, relax your jaw. Observe the sound of my voice as you become comfortable in your space". |
| **Focus on breath** | "Focus on your breath, not attempting to change it in any way, simply noticing the movement of air in and out of your body. Observe the air as it moves into your body and observe the air as it moves out of your body. As you breathe in each time, focus completely on that breath, and say to yourself "one breath." As you breathe out, focus completely on the exhale and say to yourself, "one exhale." Continue this process." |
| **Acknowledge mind wandering** | "As you continue to focus on your breath, you may find that your mind wanders; this is normal. Observe your mind wandering. Describe it by saying, "My mind wanders," and then bring your attention back to your breath." |
| **Observe thoughts** | "Observe any feelings you have in this moment, allow them to float in your mind as though waves on the ocean. As they float, describe them to yourself. For example, I feel .... However you feel in the moment is absolutely fine. Allow yourself to observe this emotion nonjudgmentally. Then gently return to the breath" |
| **Reorientation** | "As I count backward from 7 to 1, come back to the room, wiggling your fingers and toes, and moving your body gently in your chair, feeling calm yet energized 7 . . . 4, 3, 2, 1." |

