## Supplementary figures and images for "A Virtual Music Mindfulness Tool for Individuals of African descent during COVID-19"

### Supplemental Materials.pptx

#### Slide 1
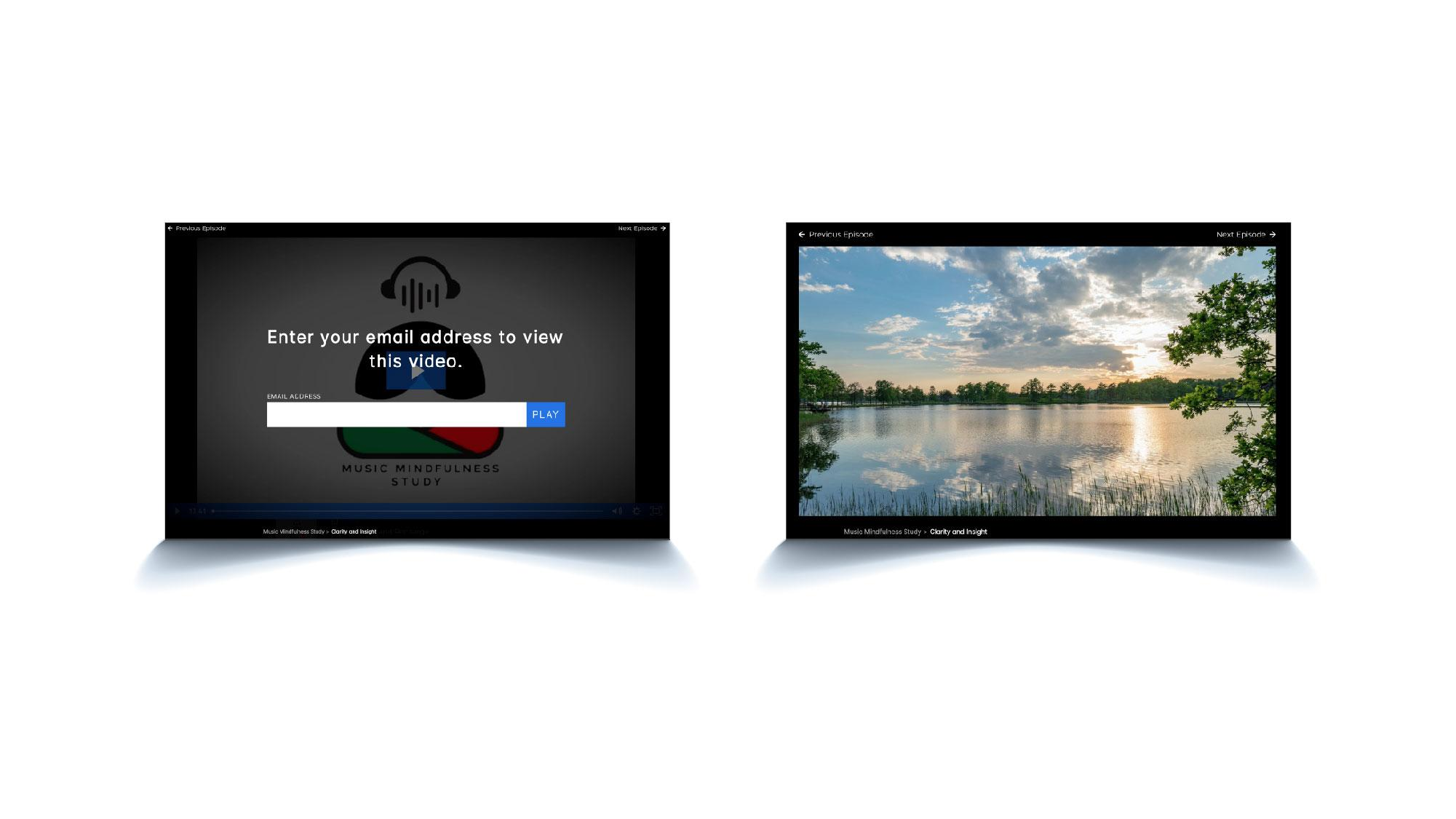

#### Slide 2
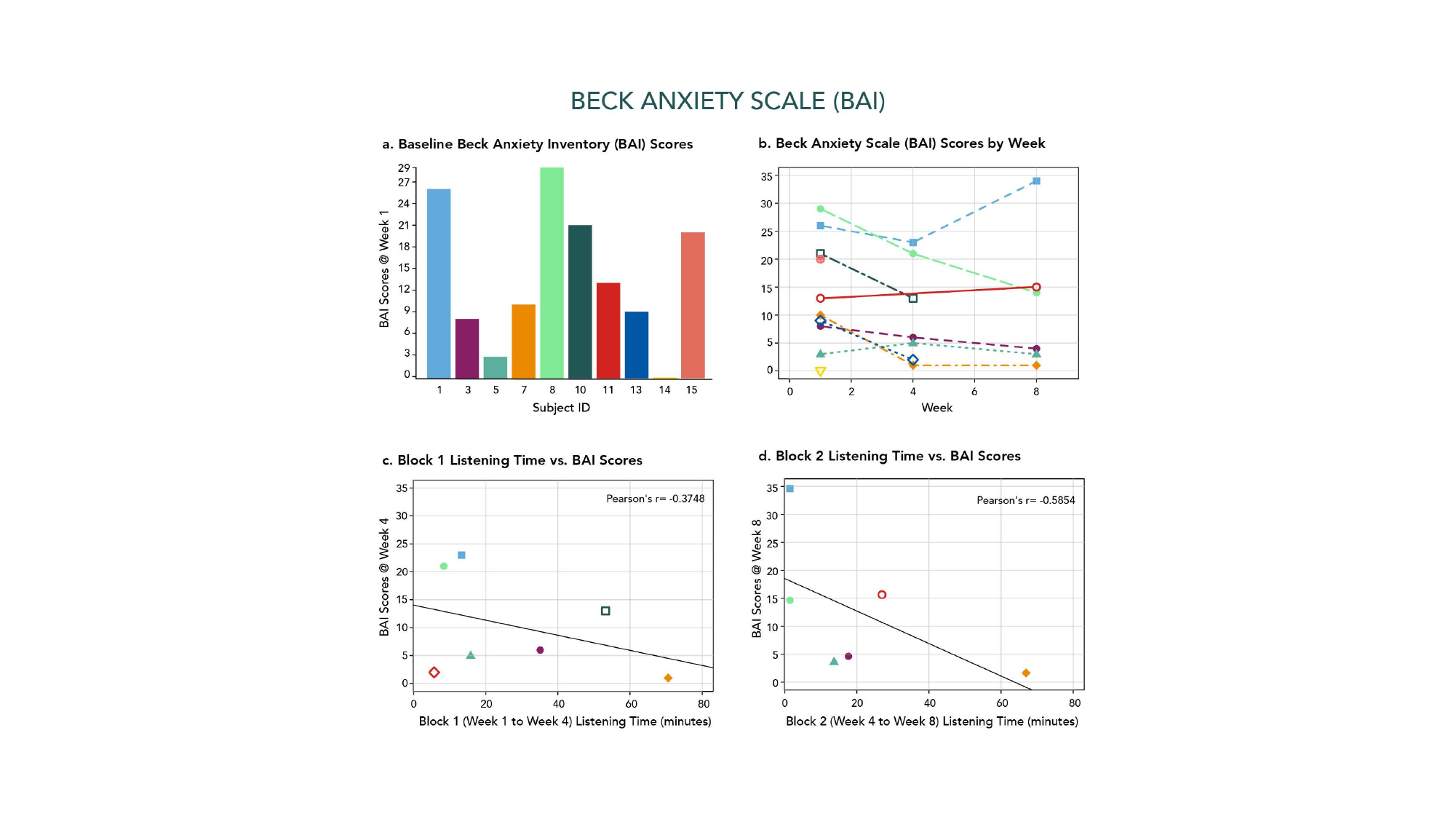

#### Slide 3
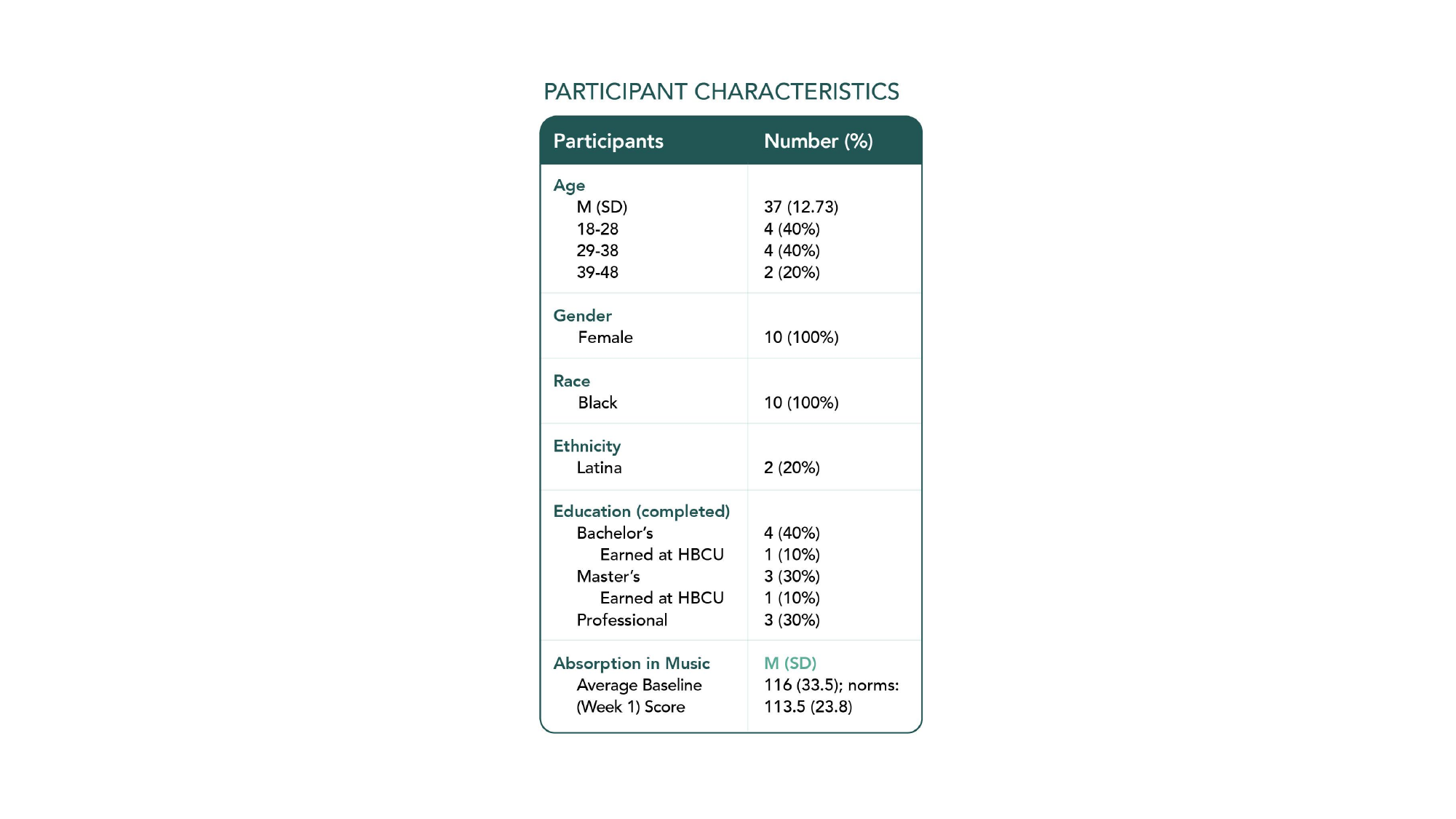

#### Slide 4
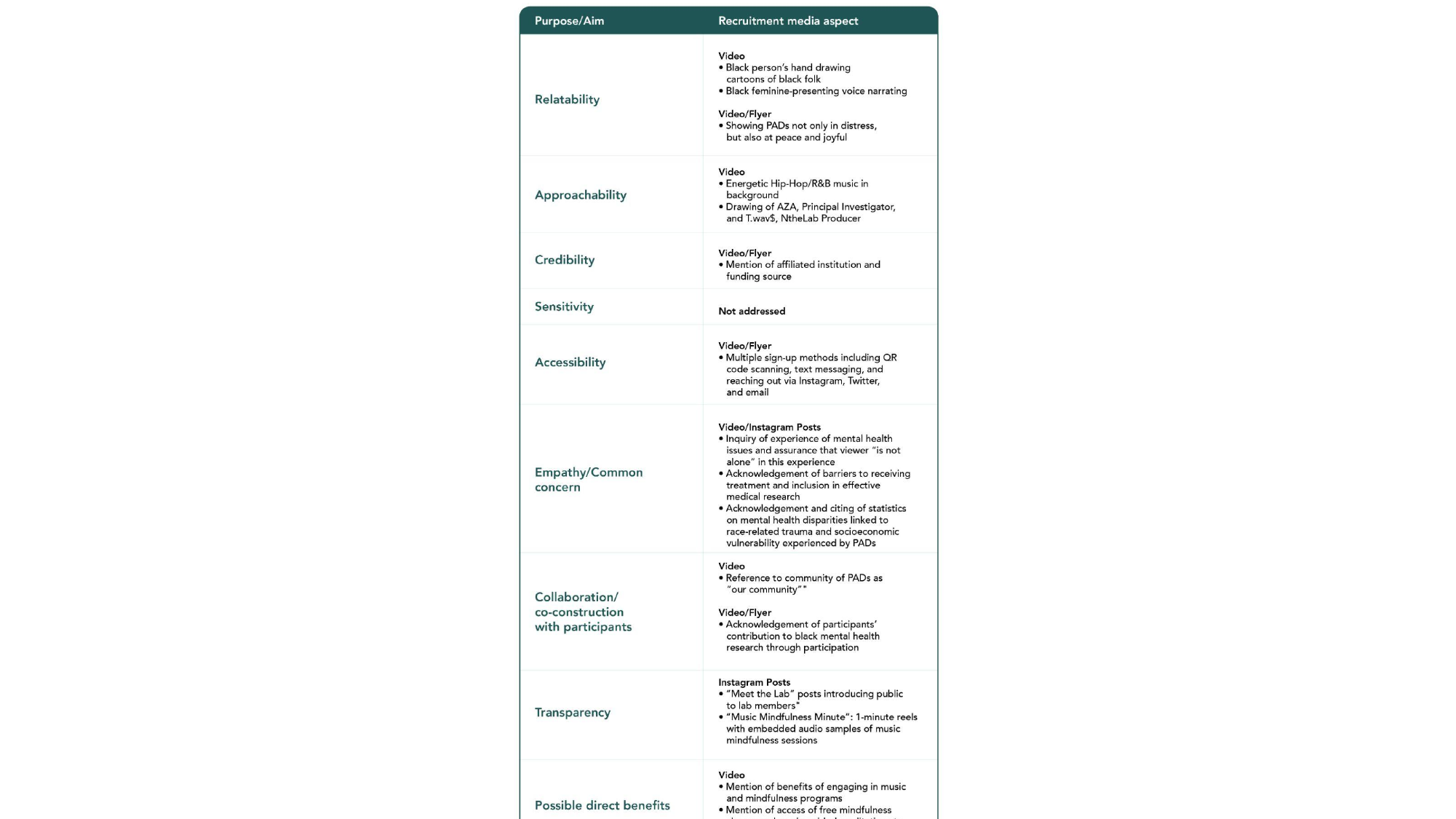

#### Slide 5
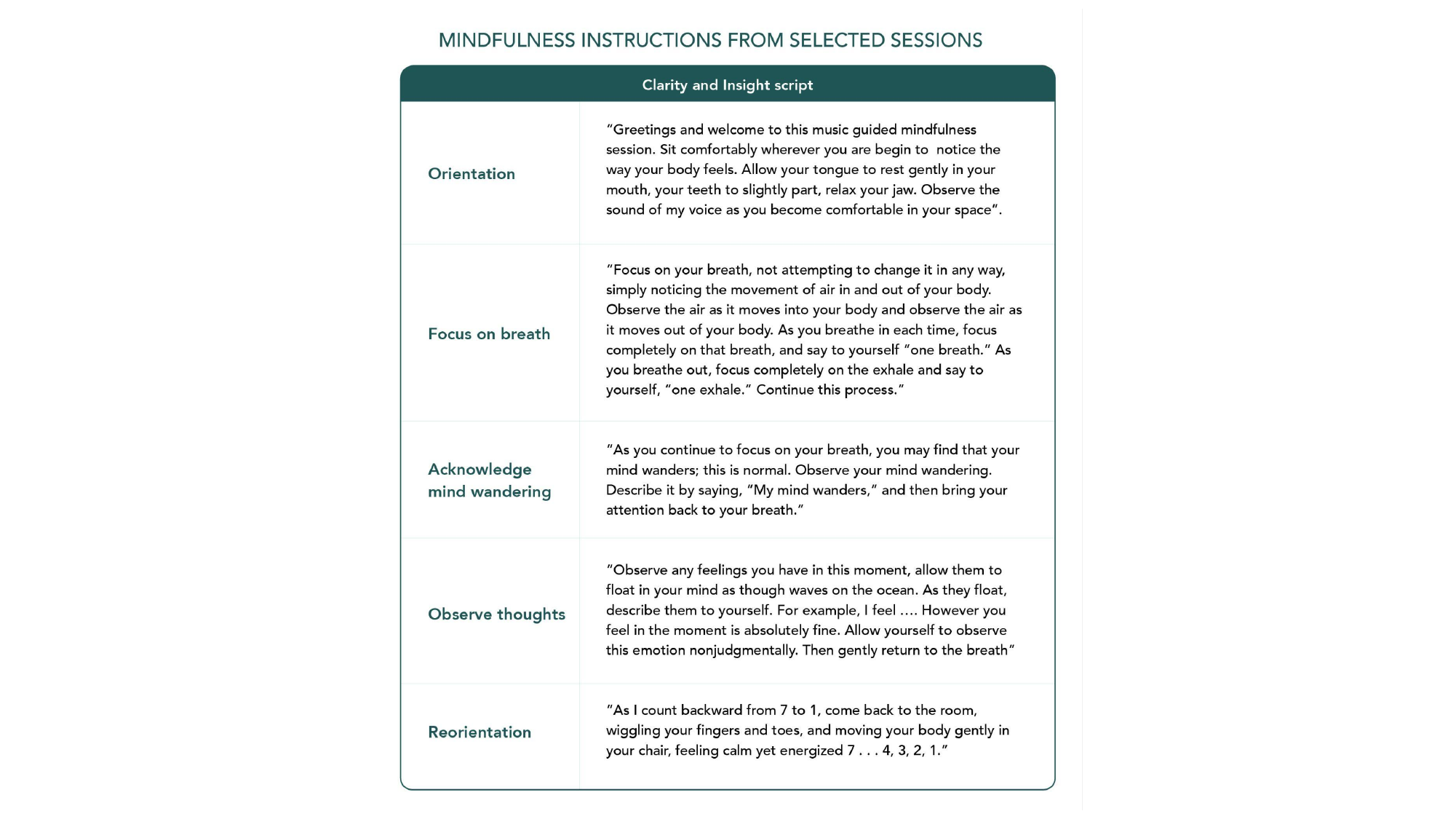
